# Validation of the Openwater wearable optical system: cerebral hemodynamic monitoring during a breath hold maneuver

**DOI:** 10.1101/2023.10.11.23296612

**Authors:** Christopher G. Favilla, Sarah Carter, Brad Hartl, Rebecca Gitlevich, Michael T. Mullen, Arjun G. Yodh, Wesley B. Baker, Soren Konecky

**Affiliations:** Department of Neurology, University of Pennsylvania, Philadelphia, USA; Open Water Internet Inc., San Francisco, USA; Department of Neurology, Temple University, Philadelphia, USA; Department of Physics and Astronomy, University of Pennsylvania, Philadelphia, USA; Department of Neurology, Children’s Hospital of Philadelphia, Philadelphia, USA

## Abstract

Bedside cerebral blood flow (CBF) monitoring has the potential to inform and improve care for acute neurologic diseases, but technical challenges limit the use of existing techniques in clinical practice. Here we validate the Openwater optical system, a novel wearable headset that uses laser speckle contrast to monitor microvascular hemodynamics. We monitored 25 healthy adults with the Openwater system and concurrent transcranial Doppler (TCD) while performing a breath-hold maneuver to increase CBF. Relative blood flow (rBF) was derived from the changes in speckle contrast, and relative blood volume (rBV) was derived from the changes in speckle average intensity. A strong correlation was observed between beat-to-beat optical rBF and TCD-measured cerebral blood flow velocity (CBFv), R=0.79; the slope of the linear fit indicates good agreement, 0.87 (95% CI:0.83-0.92). Beat-to-beat rBV and CBFv were strongly correlated, R=0.72, but as expected the two variables were not proportional; changes in rBV were smaller than CBFv changes, with linear fit slope of 0.18 (95% CI:0.17-0.19). Further, strong agreement was found between rBF and CBFv waveform morphology and related metrics. This first *in vivo* validation of the Openwater optical system highlights its potential as a cerebral hemodynamic monitor, but additional validation is needed in disease states.

## INTRODUCTION

Quantification of cerebral blood flow (CBF) at the bedside holds potential to inform and improve care for a wide range of neurologic diseases, perhaps most notably ischemic stroke in which CBF optimization is a pillar of clinical management. Unfortunately, technical limitations of existing methods for CBF quantification severely impede their clinical utility. The gold standard for non-invasive CBF imaging is O^15^-positron emission tomography (PET),^1, 2^ but O^15^-PET is logistically complicated, expensive, and exposes the patient to ionizing radiation. Advanced MRI and CT based techniques can quantify CBF, but they provide only snapshots of data and are not suitable for serial bedside monitoring.^3–7^ Invasive tissue monitors, such as the Bowman Perfusion Monitor® provide real-time physiologic data, including CBF,^8^ but they are too invasive to be practical in most clinical contexts. Thus, development and translation of a non-invasive bedside modality is needed.

Transcranial Doppler (TCD) ultrasonography is widely available and is used to serially evaluate cerebral hemodynamics in clinical practice, for example, monitoring for vasospasm after subarachnoid hemorrhage.^9, 10^ TCD is also employed to assess cerebrovascular reserve in both clinical and research settings by quantifying the change in CBF induced by a vasoactive stimulus, most commonly hypercapnia.^11, 12^ TCD provides a measure of cerebral blood flow velocity (CBFv), rather than CBF, but this limitation is mitigated by the fact that changes in velocity are proportional to changes in flow if the arterial diameter remains unchanged.^13^ Additional limitations of TCD include the requirement of a qualified technologist and the fact that nearly 20% of the population does not have adequate temporal acoustic windows, which may disproportionately affect females.^14, 15^

Another methodology, diffuse optical imaging/monitoring, is appealing because it can circumvent some of these limitations while directly assessing tissue-level physiology. Cerebral oximetry based near-infrared spectroscopy (NIRS) is widely available and often used as a surrogate of CBF.^16, 17^ However, changes in the NIRS signal may not mirror changes in CBF, *e.g.,* if there are fluctuations in arterial oxygen saturation or cerebral metabolism,^18, 19^ which is a particularly relevant limitation in cerebrovascular disease states. A qualitatively different (compared to NIRS) emerging modality is diffuse correlation spectroscopy (DCS); DCS quantifies the speckle intensity fluctuations of near-infrared light scattered by tissues to directly measure CBF.^20, 21^ DCS has been validated against gold standard O^15^-PET and other modalities,^22–26^ but signal-to-noise limitations hinder its widespread use.

In this study, we aimed to evaluate a novel, wearable optical system (Open Water Internet Inc., San Francisco CA) that illuminates tissue with short pulses of highly coherent laser light and leverages measurements of speckles and light intensity to continuously monitor microvascular hemodynamics. Like traditional DCS, the device quantifies the speckle intensity fluctuations of light scattered by tissues to measure CBF. The Openwater device, however, simultaneously samples millions of speckles via a speckle ensemble detection method that dramatically improves signal-to-noise compared to traditional DCS. The speckle analysis scheme, dubbed Speckle contrast optical spectroscopy (SCOS), has been studied by several groups,^27–32^ and typically uses a camera to measure speckle ensembles.^27–32^. A key feature of the Openwater system is its use of short pulses of very intense laser light. The use of short pulse illumination permits the dynamics of tissue deep under the surface to be probed at short time scales, while maintaining a safe low average power; thus, one need for validation stems from the use of short pulses of intense laser light, which hold potential to increase sensitivity, but which is challenging to implement without introducing spectral and modal complications that can degrade contrast. The present study utilized a 35 mm source-detector distance to measure CBF at 40 Hz sampling with sufficient SNR to resolve pulsatile CBF waveforms during the cardiac cycle. We employed a breath hold maneuver to provoke a large CBF variation in healthy volunteers to provide a means for validating the Openwater device by comparison with TCD.

## MATERIALS AND METHODS

### Participants

Healthy individuals between the ages of 18 and 45 were eligible to participate. Subjects were excluded if they had a history of hypertension, type-2 diabetes, hyperlipidemia, heart failure, stroke, cerebrovascular abnormality, intracranial mass lesion, or skull defect which could interfere with TCD monitoring at the temporal region. The study protocol was approved by the University of Pennsylvania Institutional Review Board, and all study procedures were conducted in accordance with the ethical standard of the Helsinki Declaration. All study participants provided written informed consent prior to any study procedures. The study conformed to STROBE guidelines for observational studies.

### Optical Blood Flow Instrumentation

The hemodynamic measurement device (Open Water Internet, Inc.; San Francisco, CA) consists of a wearable headset and a console. The headset contains two modules that collect data simultaneously from both side of the head. For comparison with TCD, data from the module positioned on the left lateral aspect of the forehead, overlying the lateral frontal lobe was used (**Figure 1**). The modules contain a built-in optical fiber for the delivery of low average power laser light to the surface of the brain, as well as a custom camera for the measurement of light escaping from the subject. The console contains the laser, electronics, touchscreen, and computer.

**Figure 1.**
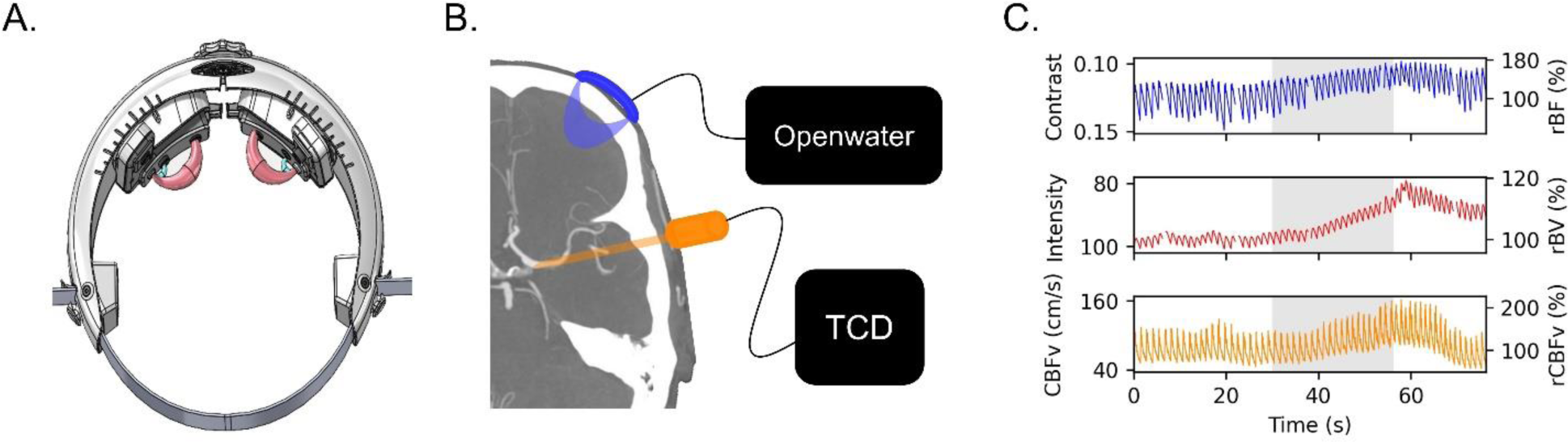
Experimental setup and raw time-series data: (A) A schematic of the Openwater headset, demonstrating the light source/detector positioning and the theoretical light path. (B) The frontal lobe is probed by the Openwater Headset over the lateral aspect of the forehead. The middle cerebral artery is insonated by transcranial Doppler. (C) An example of time-series data demonstrates one subject’s hemodynamic data during the breath hold maneuver. The blue line represents the speckle contrast (informative of flow). The red line represents the light intensity (informative of volume). The green line represents cerebral blood flow velocity as measured by transcranial Doppler. The grey shaded region represents the time during which the subject was holding their breath.

The source fibers emitted 250 µs pulses of highly coherent near-infrared laser light with wavelength near the isosbestic point for hemoglobin (785 nm). The pulses had an energy of 400 µJ and were emitted at a rate of 40 Hz. After passing through tissue, the light pulses were collected by a 3mm aperture in the module at a distance of 35 mm from the source position. Light entering the aperture passed through a bandpass filter before being detected by a 5-megapixel CMOS sensor (HM5530; Himax Technologies; Xinshi, Taiwan) optimized for NIR light. The pixel pitch of the sensor was 2 µm. The sensor was recessed from the aperture by 7 mm resulting in a coherence area (*A*_*c*_ = (*lamda* ∗ *Z*)^2^⁄*A*_*aperture*_) to pixel area (*A*_*pixel*_ = 4 *μm*^2^) ratio, *A*_*c*_⁄*A_pixel_*, of 1.1. Thus, for each camera exposure, about 5 million coherence areas (*i.e.,* speckles) were sampled. The large aperture increased light collected while only resulting in a modest decrease average speckle contrast (a 30% decrease compared to an idealized scenario where*in A*_*c*_⁄*A_pixel_* ≫ 1). The combination of a megapixel sensor and a large collection aperture, contributed to the ability of the device to make measurements at large source detector separations which would otherwise have been overwhelmed by the read noise of the sensor.

Pulsing the laser light is also a critical part of the measurement method for the following reason. To maximize our sensitivity to CBF, it is necessary to sample the speckles on the same time scale as the CBF-induced decay of the temporal auto-correlation function. This time decay scale is much shorter for multiply scattered light which samples tissue far below the surface as compared to a single scattered light reflected from the surface/near-surface tissue, as is done with laser speckle contrast imaging (LSCI). Likewise, to probe deeply, we need to maximize the separation between light source and detector. Unfortunately, collecting sufficient light over such a short period of time at a large source detector separation requires injecting high-power light (i.e., several watts) into the subject. If a continuous wave light source is used, this large average power may burn the subject. The Openwater device solves this problem by using light pulses with high peak power but at a very small duty cycle. Thus, the average power is small. The long source-detector separation (compared to the 25 mm separation used in the majority of published DCS studies) increases the depth of interrogation, and when combined with the rapid measurement scheme, increases the sensitivity to CBF changes.

For each image acquired on the CMOS sensor, the mean intensity *I* and variance *σ*^2^ were computed from the digital values of the pixels on the sensor. The variance was corrected for shot noise and read noise according, i.e., *σ*^2^ = 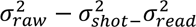. The speckle contrast (*C*) was then calculated for each image: 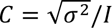. To account for other sources of variance including pixel non-uniformity and vignetting, an offset was subtracted from C. The offset corresponded to the contrast measured when the wavelength of the laser was modulated sufficiently rapidly such that its temporal coherence was reduced enough to eliminate the speckles. The resulting speckle contrast and mean intensity values were acquired at 40 Hz. We used linear interpolation to up-sample the (band limited) waveforms to 125 Hz to enable synchronization with TCD data (described below). Changes in blood flow and blood volume were estimated from changes in speckle contrast and mean intensity, respectively, as described below.

### Transcranial Doppler Ultrasonography

CBFv was assessed using a Multigon Industries® TCD system (Elmsford, NY). The left middle cerebral artery (MCA) was insonated via the trans-temporal window at a depth of 40-65 mm. The vessel was confirmed by its characteristic depth range, Doppler signal, direction, and velocity.^33^ To ensure signal stability for the duration of the monitoring period, a 2MHz TCD probe was secured directly to the Openwater Headset using a custom clamp designed to facilitate continuous vessel insonation, while minimizing motion induced artifacts or signal loss. MCA waveform (125 Hz sampling) and beat-to-beat mean CBFv were recorded and synchronized with optical data. Transient dropouts of a few TCD data points occurred, these were replaced with linearly interpolated data points.

### Cerebrovascular Reactivity Protocol

All studies were conducted in a single examination room within the Neuro-diagnostic suite at the Hospital of the University of Pennsylvania. Prior to hemodynamic monitoring, subject demographics were collected on a case report form. Skin pigmentation was assessed by the Fitzgerald scale, which quantifies skin color based on a 6-point scale. The study room was quiet and temperature controlled (23°C) throughout the duration of monitoring. Subjects were positioned in a hospital stretcher with the head-of-bed elevated to 45°. The Openwater headset (**Figure 1a**) was placed on the participant’s head to ensure the optical probes were along the upper border of the forehead (**Figure 1b**). The headset size was adjusted using a built-in dial to ensure the optical probes were on the lateral margin of the forehead (while avoiding hair). The TCD probe was secured to the Openwater headset via an adjustable clamp in order to insonate the left MCA via the temporal acoustic window (**Figure 1b**). TCD and optical data were synchronized at the beginning of each subject’s monitoring session.

After confirming signal quality from both modalities, 30 seconds of baseline data were collected. Then, a 30-second breath hold was performed. The breath hold was initiated at the end of expiration to avoid pre-oxygenation and elicit a more reliable hypercapnic response. After 2 minutes of rest, another 30-second breath hold was completed. The first breath hold was used for analysis, but if the subject was unable to perform the first breath hold or if there was signal loss with either imaging modality, then the second breath hold was used for analysis. An example of raw time series data from one subject is depicted in **Figure 1c**.

### Optical and TCD Data Processing

For each modality, a pulse-finding algorithm discriminated beats in the speckle contrast (optical) and CBFv (TCD) signals, from which beat-to-beat mean and pulsatility index [*PI* = (*peak systolic value – end diastolic value)/mean*] were obtained. A baseline value for each parameter was calculated as the average over the 30 seconds prior to initiation of the breath hold. The relative change from baseline was calculated for each beat-to-beat value thereafter (i.e., in this way, changes from baseline were effectively normalized to facilitate inter-modality comparison):

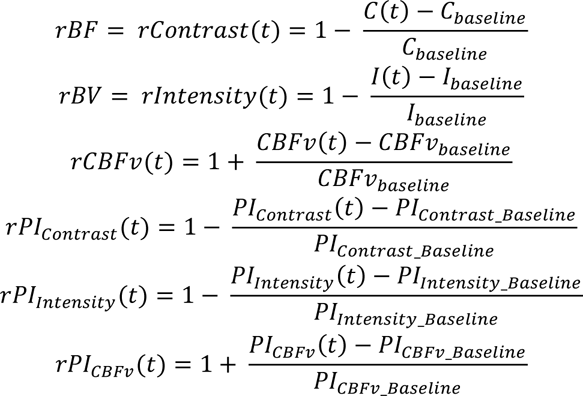

Note, we compute relative blood flow (rBF) and relative blood volume (rBV) from the fractional changes in *Contrast* and *Intensity,* respectively, during the monitoring session (*e.g.,* a 20% increase in contrast reflects a 20% decrease in blood flow; a 20% increase in intensity reflects a 20% decrease in blood volume).

Cerebrovascular reactivity was quantified by breath hold index (BHI) and time to maximum effect (i.e., seconds from breath hold initiation to the maximal value for each modality. The BHI was calculated as follows:

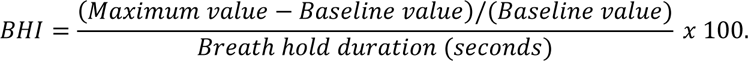

Waveform morphology was evaluated before and after the breath hold to facilitate comparison between speckle contrast-derived blood flow waveform and the TCD-derived CBFv waveform (**Figure 2**). Each pulse was normalized such that peak systolic and end diastolic values were 1 and 0, respectively. Pulses were averaged during 30 seconds of baseline data and separately averaged during the 10 second window centered at the time of maximum effect after breath hold initiation, selected at the time of peak effect post-breath hold. From these averaged pulses, a peak detection algorithm identified the dicrotic notch and three peaks: (1) P1 represents ejection of blood from the left ventricle, (2) P2 represents the pulse wave reflected by the closing aortic valve, and (3) P3 represents the diastolic flow. The augmentation index (AIx), calculated as the ratio of the amplitude of P2 to P1, provides a measure of cerebrovascular stiffness.^34, 35^ AIx was calculated based on optical blood flow (rBF-AIx) and TCD (CBFv-AIx) during baseline and hypercapnia (*i.e.,* at the end of the breath hold). Peak finding was reviewed independently by two study team members and manually corrected if necessary; notably in the pulses where 3 distinct peaks were not easily discriminated.

**Figure 2.**
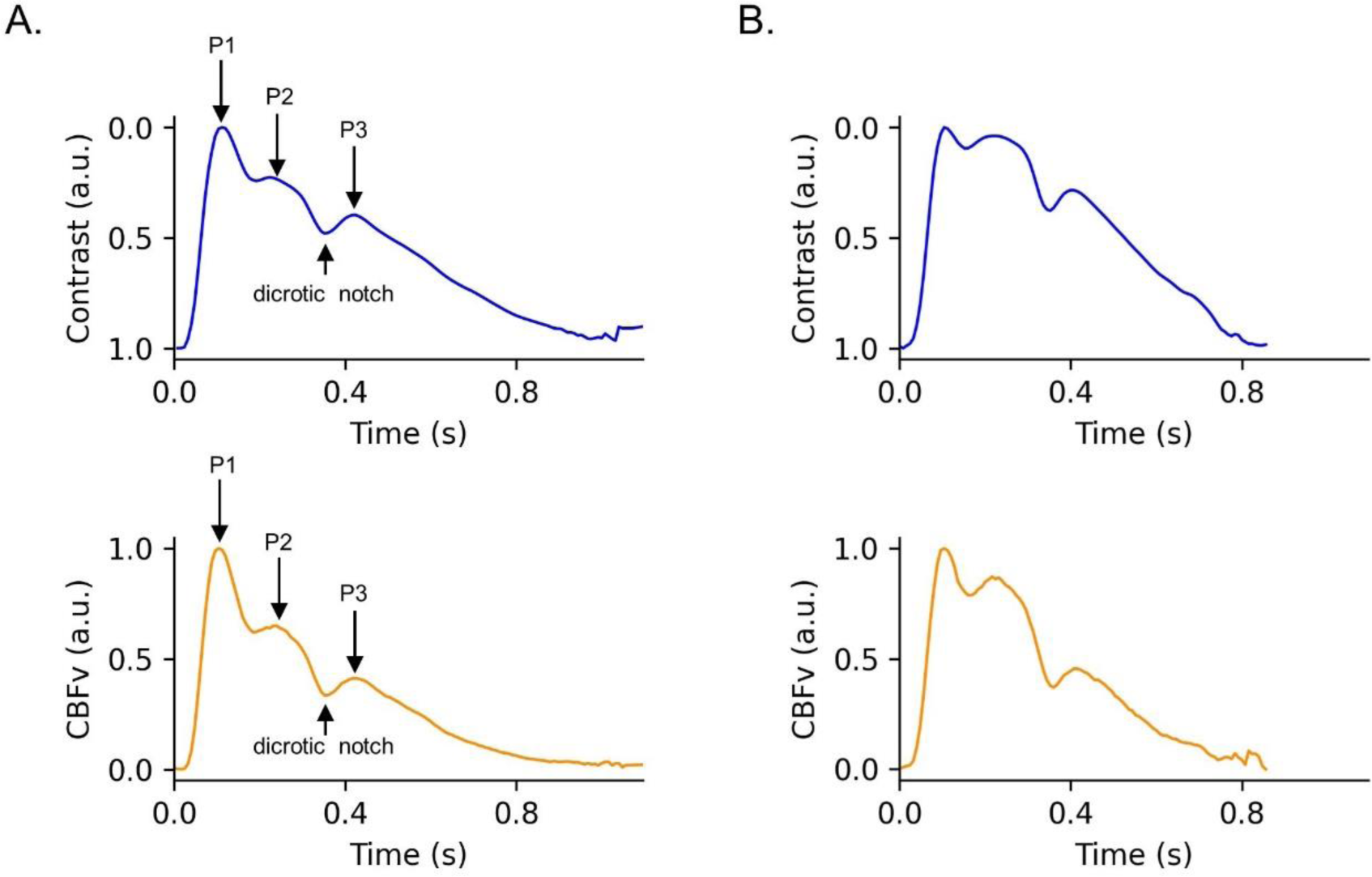
Waveform morphology before and after breath hold: The contrast waveform and CBFv waveform were averaged across 30 seconds at baseline and again across 10 seconds at the peak effect of the breath-hold. Representative waveforms are depicted from a single subject. All waveforms amplitudes are normalized (i.e. setting the y-axis scale from 0 to 1). (A) Prior to the initiation of the breath hold a single pulse wave is depicted with both modalities. The dicrotic notch and three peaks are identified (P1, P2, P3). (B) At the end of the breath hold, a change in waveform morphology, in particular an increase in the relatively amplitude of P2, can be appreciated with both modalities. CBFv indicates cerebral blood flow velocity.

### Statistical Analysis

Summary statistics were presented using means and standard deviations for continuous variables, medians and interquartile ranges for ordinal or non-parametric variables, and proportions for categorical variables. After normalizing values to the baseline period, we used correlation, mixed-effects linear regression, and Bland-Altman analyses to investigate agreement on a beat-to-beat basis between: a) mean rBF versus mean CBFv and b) mean rBV versus mean CBFv. The Pearson R was also calculated per subject. R is bounded by −1 to 1 and not expected to be normally distributed, so the average and standard deviation of R were transformed using Fisher’s transformation (F = arctanh(R), where arctanh is the hyperbolic arctangent). The resulting values were then transformed back to correlation space via the hyperbolic tangent to report summary statistics.^36^ We used Pearson’s correlation and linear regression to investigate the agreement between the optical and TCD measurements of BHI and time to maximum effect. The timing of the three peaks (P1, P2, P3) and dichrotic notch were evaluated by correlation and linear regression in comparing the optical and TCD waveforms. Finally, the pre-to post-breath hold measured change in PI and AIx were correlated between the two modalities. The data that support the reported findings are available from the corresponding author upon reasonable request.

## RESULTS

Of the 25 subjects who completed the monitoring protocol, two were excluded due to poor TCD data quality, and 23 were included in the final analysis. The first breath hold was sufficient for analysis in 21 subjects, but one subject did not correctly hold their breath on the first attempt, so the second breath hold was analyzed for this subject. The protocol was well tolerated without any adverse events. The mean participant age was 35 years (+/-11). 61% of the participants were female, and the median Fitzpatrick scale of skin pigmentation was 2 (IQR: 1 – 2).

The optical and TCD measurements of mean beat-to-beat rBF and rCBFv, respectively, demonstrated good agreement as was evidenced by a strong correlation (overall R = 0.79, R per subject = 0.88 +/-0.42) and a slope of 0.87 (95% CI: 0.83 – 0.92) in the mixed effects model (**Figure 3a**). Based on a Bland-Altman analysis, the mean difference between the two modalities was 5%, and the vast majority of the beat-to-beat values were within the 95% confidence interval for agreement (**Figure 3b**). Of additional interest was the potential agreement between beat-to-beat optical blood volume (rBV) and TCD-measured CBFv (**Figure 3c**). The correlation was strong (overall R = 0.72, R per subject = 0.85 +/-0.51), but changes in rBV were expectedly smaller than changes in rCBFv as evidenced by a slope of 0.18 (95% CI: 0.17 – 0.19) in the mixed effects model: (**Figure 3d**). The Bland-Altman analysis indicated a mean difference between the two modalities of 10% and there was a negative trend in the Bland-Altman plot because changes in rCBFv and rBV were not proportional.

**Figure 3.**
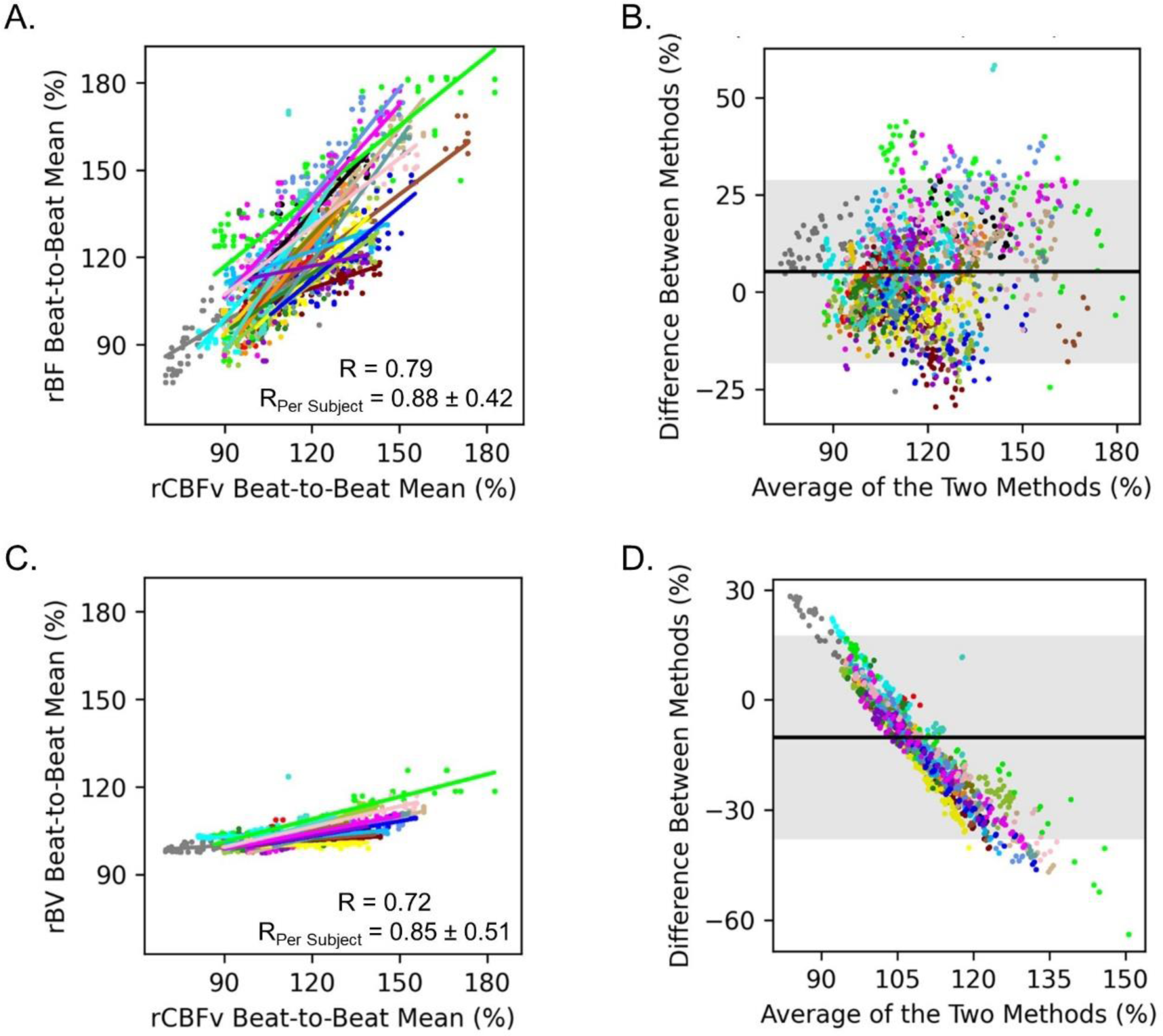
Comparing optical and TCD beat-to-beat monitoring: All data are normalized to the 30-second period preceding the breath hold. Beat-to-beat mean values are calculated for each metric from the start of the breath hold through 5 seconds after the completion of the breath hold. Each color represents a different subject. (A) A scatterplot depicts the beat-to-beat mean rCBFv (x-axis) and the beat-to-beat mean rBF (y-axis). The overall correlation coefficient is 0.79. The average correlation coefficient (when calculated for each subject individually) is 0.88 (+/-0.42). The slope of the mixed-effects linear model is 0.87 (95% CI: 0.83 – 0.92). (B) A Bland-Altman plot indicates beat-to-beat mean rCBFv is on average 5% smaller than beat-to-beat mean rBF. The grey shaded region represents the 95% confidence interval for agreement. (C) A scatterplot depicts the beat-to-beat mean rCBFv (x-axis) and the beat-to-beat mean rBV (y-axis). The overall correlation coefficient is 0.72. The average correlation coefficient (when calculated for each subject individually) is 0.85 (+/-0.51). The slope of the mixed-effects linear model is 0.18 (95% CI: 0.17 – 0.19) which indicates that changes in rBV are smaller than changes in rCBFv. (D) A Bland-Altman plot indicates rCBFv is on average 10% larger than rBV. The grey shaded region represents the 95% confidence interval for agreement. A negative trend is evident and indicates that as the average value increases, the difference between CBFv and rBV increases. TCD indicates transcranial Doppler. rCBFv indicates TCD measured relative cerebral blood flow velocity. rBF indicates optically measured relative blood flow. rBV indicates optically measured relative blood volume.

There was good agreement observed between the BHI calculated based on optically measured blood flow and TCD measured CBFv (**Figure 4a**). The correlation coefficient was 0.78 and the slope of the line of the best fit was 0.85 (95% CI: 0.54 – 1.16). There was also a strong correlation between BHI values calculated based on blood volume and CBFv (**Figure 4b**; R = 0.75), but again rBV-based BHI values were expectedly smaller (slope = 0.22; 95%CI: 0.13 – 0.31). The time from breath hold initiation to maximum cerebral hemodynamic effect was also compared across modalities (**Figure 5**), and there was strong agreement between the rBF and rCBFv timing [R = 0.92, slope = 0.90 (95% CI: 0.72 – 1.08)], as well as the rBV and rCBFv timing [R = 0.92, slope = 0.91 (95% CI: 0.74 – 1.08)].

**Figure 4.**
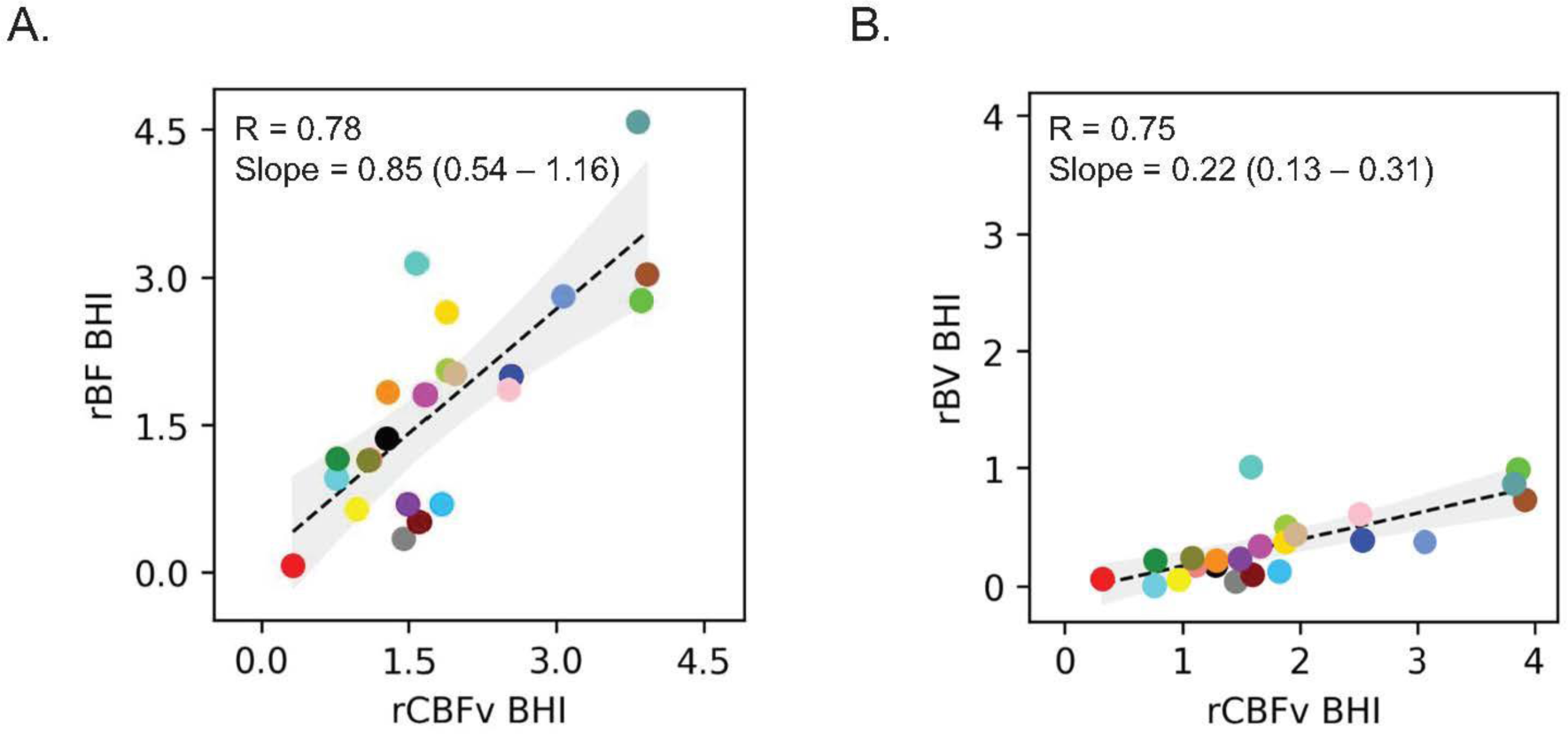
Calculating breath hold index with optics and TCD: The breath hold index was calculated for each metric. (A) A scatterplot depicts the BHI based on TCD-derived CBFv (x-axis) and the BHI based on optically-derived rBF (y-axis). The correlation coefficient is 0.78. The linear regression coefficient is 0.85 (95% CI: 0.54 – 1.16). (B) A scatterplot depicts the BHI based on TCD-derived CBFv (x-axis) and the BHI based on optically-derived rBV (y-axis). The correlation coefficient is 0.75. The linear regression coefficient is 0.22 (95% CI: 0.13 – 0.31). TCD indicates transcranial Doppler. rBF indicates optically measured relative blood flow. rBV indicates optically measured relative blood volume. rCBV indicates TCD measured relative cerebral blood flow velocity. BHI indicates breath hold index.

**Figure 5.**
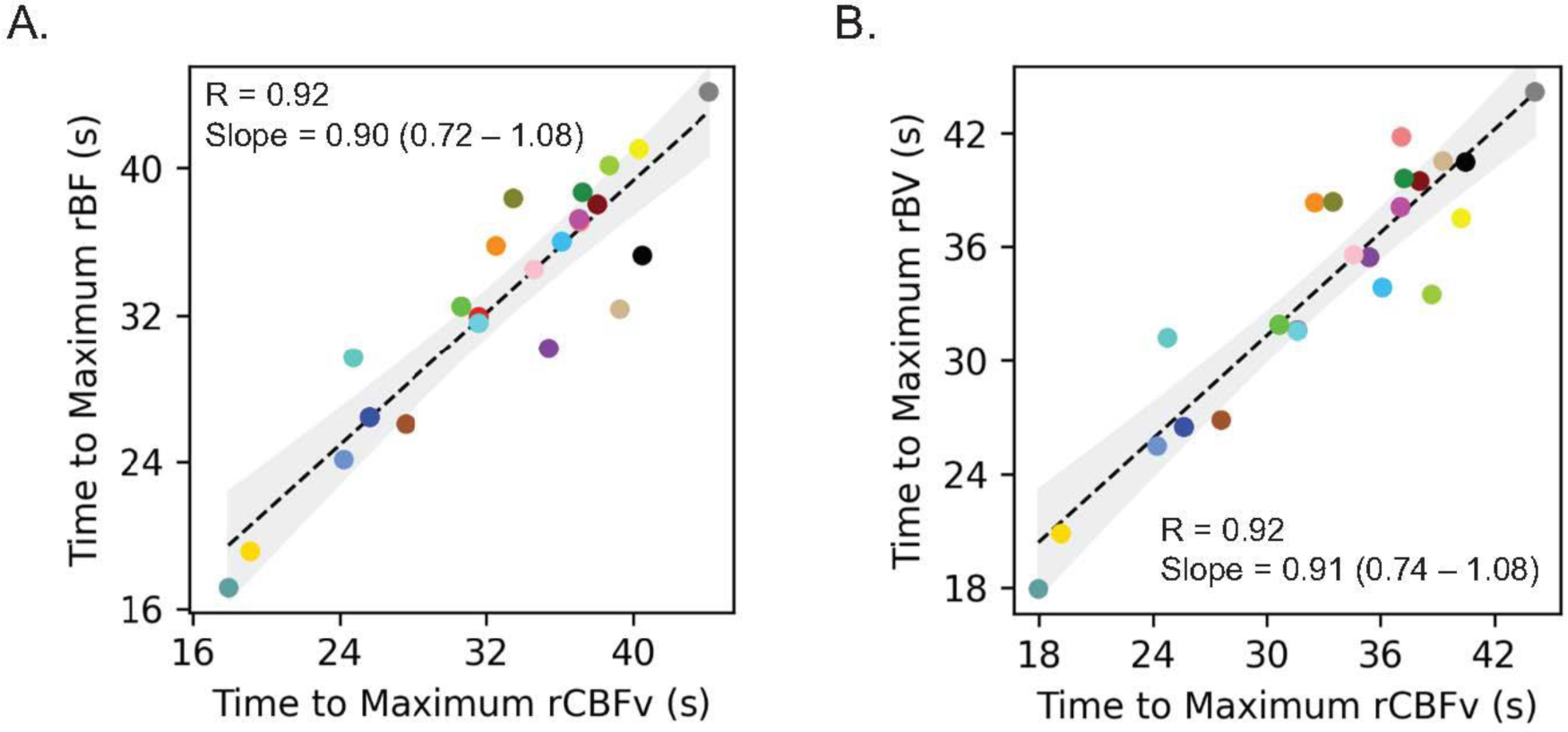
Timing of the cerebral hemodynamic effect: Time (seconds) was calculated from the initiation of the breath hold to the maximum effect for each metric. (A) A scatterplot depicts the time to maximum effect for rCBFv (x-axis) and for rBF (yaxis). The correlation coefficient is 0.92. The linear regression coefficient is 0.90 (95% CI: 0.72 – 1.08). (A) A scatterplot depicts the time to maximum effect for rCBFv (x-axis) and for rBV (yaxis). The correlation coefficient is 0.92. The linear regression coefficient is 0.91 (95% CI: 0.74 – 1.08). rBF indicates optically measured relative blood flow. rBV indicates optically measured relatively blood volume. rCBV indicates relative cerebral blood flow velocity. s indicates seconds.

We finally compared the timing of morphologic features of the rBF and rCBFv waveform (*i.e.,* P1, P2, P3 and the dichrotic notch; see **Figure 2**). There was good agreement between the two modalities with respect to peak timing within the pulse, based on correlation and slope of the best fit line for each peak (**Figure 6a**). There was similarly good agreement between the two modalities with respect to the timing of the dicrotic notch within the pulse, with a correlation coefficient of 0.84 and a slope of 0.70 (95% CI: 0.50 – 0.91) for the best fit line (**Figure 6b**).

**Figure 6.**
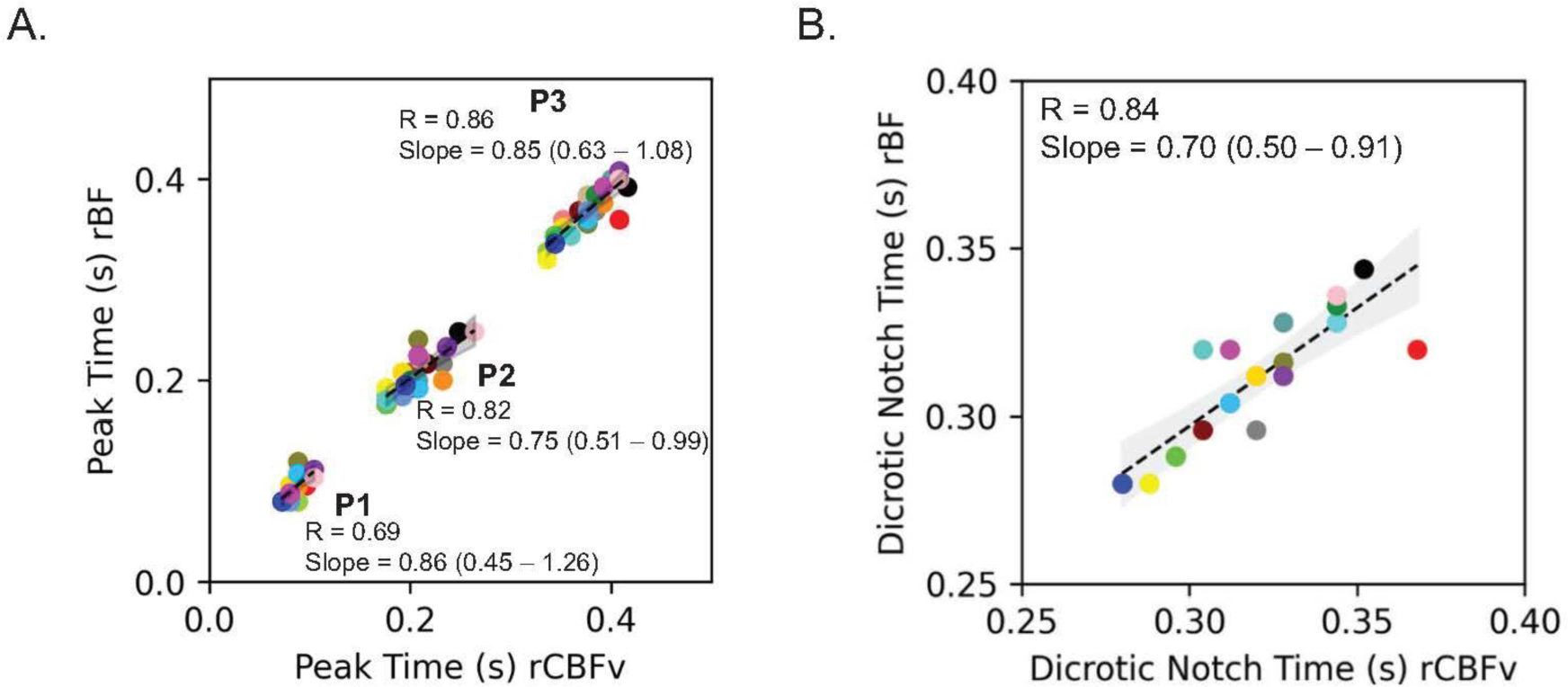
Timing of waveform features: For each subject waveforms were averaged across the 30 second baseline period. A peak-fiinding algorhythm identified the dicrotic notch, P1, P2, and P3. (A) A scatterplot depicts the timing of each peak based on rCBFv (x-axis) and rBF (y-axis). The correlation coefficient for P1 is 0.69, and the linear regression coefficient is 0.86 (95% CI: 0.63 – 1.08). The correlation coefficient for P2 is 0.82, and the linear regression coefficient is 0.75 (95% CI: 0.51 – 0.99). The correlation coefficient for P3 is 0.86, and the linear regression coefficient is 0.85 (95% CI: 0.45 – 1.26). (B) A scatterplot depicts the timing of the dicrotic notch based on CBFv (x-axis) and rBF (y-axis). The correlation coefficient is 0.84, and the linear regression coefficient is 0.70 (95% CI: 0.50 – 0.91). rBF indicates optically measured relative blood flow. rCBV indicates relative cerebral blood flow velocity. s indicates seconds.

Of further note, an expected reduction in PI was observed during hypercapnia as the pulse width became reduced, and this effect was very strongly correlated between the two modalities (R = 0.84; **Figure 7a**). Similarly, an expected increase in AIx observed during hypercapnia as the amplitude of P2 increased, and this change was strongly correlated between the two modalities (**Figure 7b**).

**Figure 7.**
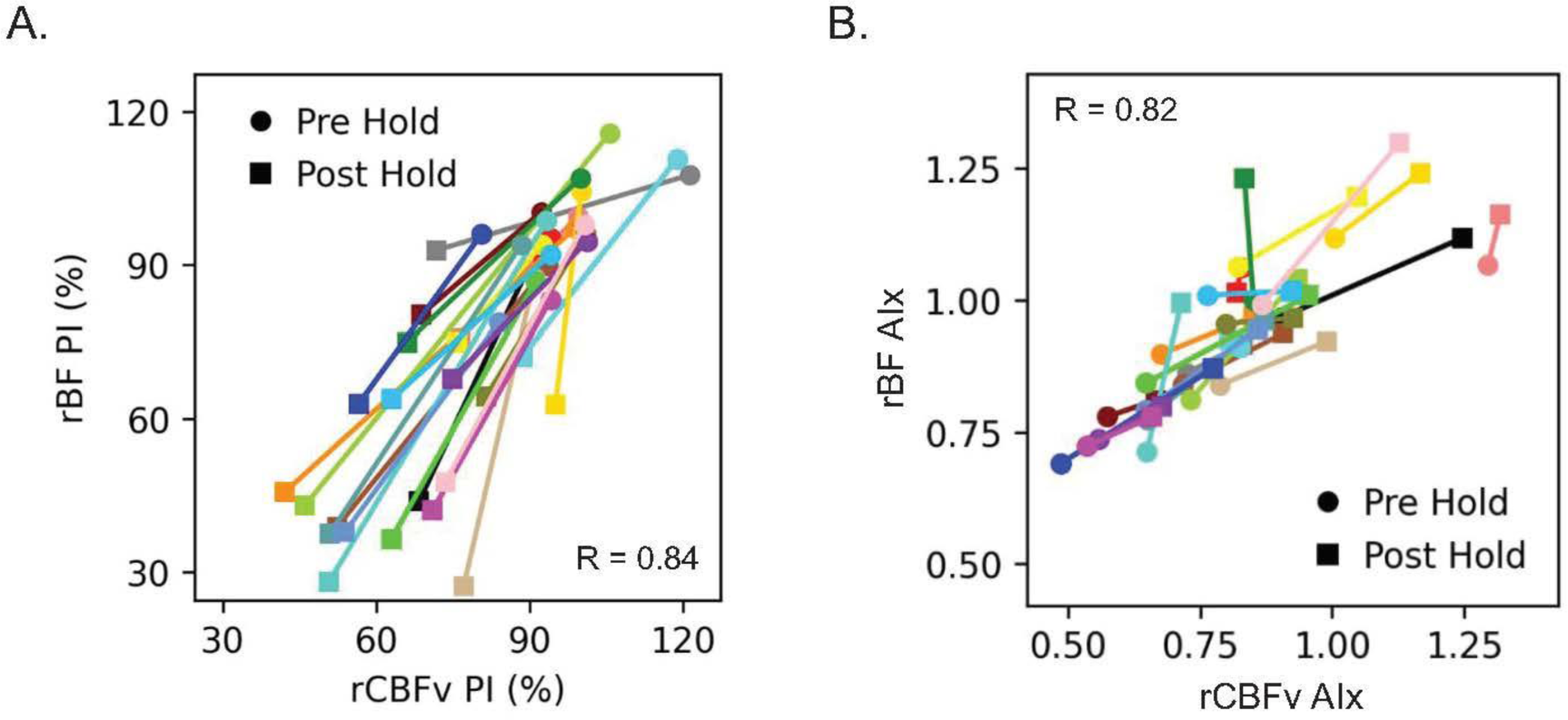
Change in PI and AIx during breath hold: (A) A scatterplot depicts PI based on rCBFv (x-axis) and rBF (y-axis). Each subject has a data point pre-hold and post-hold. PI is smaller post-hold because the pulse pressure is reduced during hypercapnia. The correlation coefficient is 0.84. (B) A scatterplot depicts the AIx (i.e. P2/P1) based on rCBFv (x-axis) and rBF (y-axis). Each subject has a data point pre-hold and post-hold, and the AIx is larger post-hold which reflects a relative increase in the P2 amplitude. The correlation coefficient is 0.82. rBF indicates optically measured relative blood flow. rCBFv indicates relative cerebral blood flow velocity. PI indicates pulsatility index. AIx indicates augmentation index.

## DISCUSSION

The Openwater Headset is a promising non-invasive optical system that can be leveraged to monitor cerebral hemodynamics at the bedside. This study is the first in-vivo validation of the Openwater system. Validation was obtained by comparing changes in optically-derived metrics with changes in TCD-metrics during a breath hold maneuver. Changes in the speckle contrast reflect changes in blood flow and were shown to strongly correlate with TCD at the beat-to-beat level. The breath hold index measures the overall change in CBF associated with breath hold and can be calculated with both speckle contrast and TCD with good agreement. Although TCD flow velocity correlated with optical blood volume, as expected, the changes were not proportional because cerebral blood volume pulsatility during the cardiac cycle and blood volume responses to hypercapnia are smaller than the corresponding flow changes.^37–39^ Importantly, high frequency data collection with the Openwater Headset allows for characterization of blood flow waveform morphology. Openwater and TCD measurement of peak times and clinically useful waveform-based metrics, such as PI and a vascular stiffness index, were very strongly correlated. This initial validation of the Openwater Headset motivates justifies future validation in larger cohorts and in clinically relevant disease states such as stroke.

The correlation between rBF and rCBFv is particularly noteworthy because it was observed not just for steady-state changes during breath-holding, but also for individual beat-to-beat changes. Although the 95% confidence interval of agreement was relatively broad, beat-to-beat values are particularly sensitive to movement artifacts or changes in signal quality over the course of monitoring. Still, the beat-to-beat correlation for each subject was strong. Any subject-level variability that exists is not explained here but could be addressed in a larger cohort with attention to potential contributions from skin pigmentation, age, or skull thickness. In future work, comparison with additional modalities such as O^15^-PET or ASL-MRI would provide further validation in a different experimental model.

Change in blood volume was expected to be smaller than the change in flow.^37–39^ The relationship between flow and volume can be summarized by the central volume principle (CBV = CBF mean transit time).^40^ With hypercapnia, as flow increases, there is an observed reduction in transit time which indicates an increase in venous drainage, thereby blunting the increase in volume.^39^ Cerebral blood volume is a key contributor to intracranial pressure, so blunting the increase in blood volume helps to avoid a potentially catastrophic increase in intracranial pressure.^41^ Alternatively, the increase in blood volume may be counterbalanced by displacement of cerebrospinal fluid in order to minimize the effect on intracranial pressure.^42^ In clinical settings, quantifying both rBF and rBV is useful because the combination provides a more thorough characterization of cerebral hemodynamics; e.g., the ratio of flow and volume is informative of transit time and regional perfusion pressure,^39, 43^ which has implications across a range of neurologic disorders including ischemic stroke, hemorrhagic stroke, subarachnoid hemorrhage, and traumatic brain injury. There are potential systematic error in quantifying blood volume. Changes in oxygenation could impact intensity without a change in blood volume, but wavelength of the 785 nm laser is very close to the isosbestic point of hemoglobin, so even large changes in oxygenation are expected to result in very small changes in intensity.^44^ Further, at 785 nm, any difference in the overall absorption of hemoglobin should be very small relative to the isosbestic point. Hypercapnia may briefly impact pH which in turn may affect the hemoglobin absorption spectrum, but this effect is expected to be very small at the end of the breath-hold.^45^

The high frequency data acquisition allows the Openwater Headset to discern several morphologic features of the blood flow waveform (speckle contrast). Visualizing the expected peaks and dicrotic notch within the rBF waveform provides an important degrees of face validity, and the strong agreement between morphologic features between optical and TCD waveforms is reflective of both construct and content validity. Finally, the correlation between dynamics of PI and AIx provides criterion validity. The optical technique described here has sufficient SNR to resolve the CBF waveform because of its use of short pulses of intense light, unlike continuous wave speckle contrast optical spectroscopy as has previously been reported in the literature.^29, 30^ In a fiber-based SCOS system, Kim et al. observed an improvement in SNR and blood flow waveform detection using a rotating chopper wheel to pulse the light.^32^ The Openwater system uses shorter pulse lengths (250μs rather than >2ms) which allow dynamics to be probed at a shorter time scale, thereby increasing the sensitivity of contrast to small changes in flow and improving waveform detection. Contrast measured using shorter pulses are also effectively more sensitive to longer photon pathlengths, i.e., pathlengths that are more biased toward brain than scalp. In practice, using short intense pulses is technically challenging. Short-pulse high peak power laser operation can lead to chirping which degrades coherence, and high power laser amplification can result in multiple fluctuating spatial modes that can also reduce signal-to-noise ratio. However, our study shows explicitly that these potential complications were not significant (at least for the present measurements at 35mm source-detector separations on the forehead). The Openwater system is also uniquely designed to include the cameras within the headset (rather than fiber-based headsets), thus ameliorating motion artifacts. The Openwater Headset’s small portable design improves convenience in certain clinical applications. Diffuse correlation spectroscopy has been used to quantify waveform features^46–48^ but with lower SNR.^32^

In clinical practice, CBF waveforms are expected to be informative of cerebrovascular resistance, compliance, and intracranial pressure.^48–50^ TCD-derived CBFv waveform is often interpreted to that end,^51^ but a low-cost user-friendly optical system may have distinct advantages as it evades the need for a trained ultrasonographer and is not limited to patients with adequate temporal acoustic windows. Data in patients with abnormal cerebral hemodynamics would contribute to instrument validation and would help to assess feasibility in an eventual clinical application. For example, a hallmark of acute stroke care is optimization of CBF, but CBF is rarely measured in practice, so there is an opportunity to apply a bedside hemodynamic monitor to facilitate physiology-guided care. As previously described, the ability to measure the blood flow waveform may prove useful, but further study is needed to determine if the optical waveform morphology is informative of clinically relevant pathology, such as elevated intracranial pressure or impaired cerebrovascular compliance.^51, 52^ In acute stroke patients, the TCD waveform may have a role in detection of large vessel occlusions,^53^ but this has not yet been described with biomedical optics.

Despite the encouraging results, this study has several important limitations. Generalizability is limited because of its small numbers and its relatively narrow range of ages and skin pigmentation. Darker skin pigments absorb more light,^54^ so it is critical to demonstrate that the agreement reported here is not pigment-dependent. No test-retest analysis was performed to assess intra-rater reliability because breath holding is often inconstant, but a future study could use a more reproducible change in CBF to evaluate test-retest reliability. Using TCD as the comparator is noteworthy because it provides a measure of CBFv rather than CBF. However, in this experimental model changes are monitored over a very short period of time during which the MCA trunk diameter is expected to remain stable, so relative changes in TCD are reflective of changes in CBF. Further, TCD is commonly used to calculate the BHI in clinical practice. The degrees of hypercapnia was not quantified in each subject, which may appear to be a shortcoming, but in actuality the precise change in PaCO_2_ is not relevant to the validation because both modalities were observing the same change in CBF. However, if any subjects had a very small change in CBF, it may have been helpful to know if those subjects had a very small change in PaCO_2_.

The Openwater system is a promising non-invasive laser speckle-based cerebral hemodynamic monitor. The compact design facilitates portability and the simple user interface emphasizes the potential for future clinical translation. This system’s first *in vivo* validation was demonstrated herein via comparison to TCD. Several data elements were scrutinized to allow for a more robust validation: (1) beat-to-beat changes, (2) breath hold index, (3) waveform morphology, and (4) dynamics of waveform-based metrics. In total, these analyses are encouraging of future work aimed at validating the Openwater system in disease states, such as stroke, in which a significant need for a practical bedside cerebral hemodynamic monitor exists.

## Data Availability

The data that support the reported findings are available from the corresponding author upon reasonable request.

## Acknowledgements

The authors greatly appreciate the technical and engineering support provided by Peter Herzlinger.

## Author contributions

CGF was responsible for study design, data acquisition, data interpretation, and manuscript preparation. SC and BG were responsible for and data acquisition, database management, and manuscript revisions. BH was responsible for instrumentation, data processing and analysis, and manuscript revisions. MTM was responsible for study design, data interpretation, and manuscript revisions. AGY was responsible for study design, data interpretation, and manuscript revision. WBB was responsible for study design, data interpretation, and manuscript revisions. SK was responsible for instrumentation, data processing, data interpretation, and manuscript revisions.

